# Overview of maternal morbidity in Morocco Marrakech-Safi region

**DOI:** 10.1101/2022.09.27.22280423

**Authors:** Hanane Hababa, Bouchra Assarag

**Affiliations:** Regional Directorate of Health and Social Protection Marrakech-Safi Morocco; National School of Public Health Rabat Morocco

**Keywords:** Maternal morbidity less severe, measurement tools, antenatal care, postpartum care, maternal and child health

## Abstract

**Objectives:** The measurement of less severe maternal morbidity represents many challenges for women during pregnancy and postpartum, and for the organization of health services. This article aims to test the tools for measuring maternal morbidity less severe proposed by the WHO and describe the state of play of this morbidity at the level of the prefecture of Marrakech. The second phase is to study the feasibility and acceptability of integrating these tools into the Pregnancy and Childbirth Surveillance Program.

**Methods:** The analysis focused on the maternal morbidities declared and diagnosed in the test, and related the feasibility and acceptability as well as the relevance of the tools tested.

**Results:** Most of the women who participated in the study (55.95% antenatal and 52.17% postpartum care) were not in good health (A medical or obstetric condition is diagnosed). Of these women, 35.79% had direct (obstetric) complications and 33.85% indirect (medical) complications. In terms of feasibility, the results suggest that the implementation of the tools presents challenges in terms of time, resources and coordination. Regarding the acceptability of the WMOs, the women surveyed perceive it as a useful information tool that promotes communication with health professionals and makes it possible to assess their state of health and ensure their holistic care.

**Conclusion:** Considering these results, the approach supports the relevance of implementing the MM measure in antenatal and postpartum care to improve the quality of care for women, to promote communication and continuity of care. However, constraints of time, resources and coordination must be taken into account for its implementation at primary health care.

## Introduction

Maternal health is critical to sustainable development and is a serious problem worldwide. The magnitude of this problem puts maternal well-being and survival at the forefront of all countries [2]. Each year, approximately 210 million women become pregnant and about 140 million newborns are born which means that maternal health is not a marginal issue [3]. In 2015, WHO estimated that 303,000 women died from pregnancy or childbirth-related complications, most of these deaths were preventable. Given this alarming situation, there is greater global awareness of the plight of women who have pregnancy or childbirth complications and may continue to have long-term problems [4]. Fortunately, women’s health and their abilities to perform economic and social functions are a central concept of the Sustainable Development Goals (SDGs). For example, the “Survive, Thrive and Transform” agenda of the Global Strategy for Women’s, Children’s and Adolescents’ Health (2016-2030), moves away from the goal regarding the reduction of maternal and child mortality, emphasizing the need to ensure good health so that women, adolescents and children can play their full role in future development.

According to Lale Say et al [1], maternal mortality is only part of the overall burden of maternal ill health, as it excludes maternal morbidity. The burden of maternal morbidity is not yet known [5]. WHO estimates that for every recorded maternal death, 20 to 30 women suffer morbidity. Of these cases, one quarter may suffer severe and permanent sequelae. Maternal morbidity, represented by the health problems suffered by women during pregnancy, delivery and the postpartum period, contributes to this burden [6]. According to the same authors, these sequelae can affect women physically, mentally, sexually, in their ability to function (cognition, mobility, participation in society), body image and socioeconomic status. Not surprisingly, the burden of both maternal morbidity and mortality is estimated to be highest in low- and middle-income countries, particularly among the poorest women. Currently, there are an estimated 27 million episodes of direct complications that occur each year [1].

Despite this large number of complications, there is little in-depth research on maternal morbidity. The range of conditions is so broad that studies often focus on the most serious and lethal causes of obstetric morbidity and/or a single disease [2].

In 1999, Fortney and Smith [7] noted that the literature is replete with hospital-based, case-based studies describing acute and chronic morbidities related to pregnancy and childbirth. What remains relatively unknown is the prevalence of specific or general morbidity in the general population. These remarks remain largely valid today, with little morbidity data having changed, despite the increase in maternal health data [8, 9].

In Morocco, the attention given to maternal health in recent years has resulted in significant progress in improving maternal and neonatal health. Considerable progress in reducing maternal and neonatal mortality has been noted. Indeed, according to the latest national survey conducted by the Ministry of Health in 2017 and 2018, the maternal mortality rate is estimated at 72.6 maternal deaths per 100,000 live births with a decline of nearly 78% compared to the 1992 NPHS. The neonatal mortality rate declined by 38% between 2011 and 2018, from 21.7 to 13.56 per 1000 NV. The improvement in these health care indicators as evidenced by the survey results shows that Morocco is on track to achieve the SDGs. While the current focus is on survival during pregnancy and childbirth, and on ensuring that women thrive throughout their lives, much remains to be done to make pregnancy a positive experience. In order to meet Sustainable Development Goal 3 and the global strategy, Morocco is called upon to develop integrated health services that maximize women’s health, well-being, and potential throughout the life cycle. In building the new national maternal and newborn health strategy to end preventable deaths, improving maternal health measurement will be key to success [10].

Building on WHO’s success in defining and measuring severe maternal morbidity (Near miss), the focus is now on standardizing and measuring less severe maternal morbidity. Indeed, in 2012, WHO [11] launched a work program on the development of a common definition and identification criteria for very severe maternal morbidity (Near-miss) allowing its routine measurement and monitoring, in particular as a tool for assessing the quality of care for women with severe morbidity. However, the definition and criteria did not exist for less severe cases along the maternal health continuum. This working group agreed on the following definition of maternal morbidity: “any health condition that contributes to and/or complicates pregnancy and childbirth and has a negative impact on the woman’s well-being and/or functioning. This work led to a conceptual framework, entitled Maternal Morbidity Measurement (MMM) Framework [12].

To address the need to measure and respond to the magnitude of this burden, a maternal morbidity measurement tool (MMT) has been developed. This tool is unique in that it is regionally sensitive and adaptable to any community category. It measures less severe maternal morbidity in prenatal and postnatal clinical populations. This tool has already been tested in three resource-limited countries, namely Jamaica, Kenya, and Malawi [13]. No tests have been conducted to date in the Arab or North African countries. Therefore, the adoption of maternal morbidity measurement tools in our Moroccan context requires a study to determine the feasibility and acceptability of setting up a simple and adapted tool.

In this sense, the present study is proposed to describe the burden of less severe maternal morbidity in the city of Marrakech, and to test the data collection tool proposed by the WHO in order to arrive at a standard tool, adapted to our context. The data collected through this study will provide better information on the extent of maternal morbidity.

In order to address the current gaps in maternal and newborn health care, this study will equip researchers, policy makers, and health care professionals to plan interventions and resources to help meet women’s reproductive health needs. This study will test the feasibility and acceptability of using a tool to measure the impact of pregnancy and childbirth on women’s health status. Once validated in our context, this WMO will provide evidence and will be a routine tool for measuring maternal morbidity.

## Framework

This conceptual framework, developed by the WHO Maternal Morbidity Working Group, highlights the broad ramifications of maternal morbidity and outlines the types of measures needed to capture what is relevant to women, service providers, and policymakers. This new conceptual framework presents a new definition of maternal morbidity and its measurement. It takes into account and applies the principles of the WHO International Classification of Functioning, Disability and Health (ICF) [14].

### Objective of the study

The objective of this study is to describe the current state of maternal morbidity in the prefecture of Marrakech.

## Method

The present study concerned pregnant women whose pregnancy was 28 weeks and older, followed up at one of the ten health centers where the study was conducted, as well as women who had given birth six weeks earlier, followed up in post-natal consultations at the same centers. Referring to the research protocol already proposed by the WHO [1], the morbidity measurement tools were tested on more than 510 women (257 for the prenatal period and 253 for the postnatal period). We adopted this sample to have the possibility of comparing with the countries where this study was conducted (Jamaica, Malawi and Kenya). Women were recruited for ANC during the period of data collection and for late PNC or for vaccination of her child. These two measurement tools were translated from English to French, validated by resource persons and tested. Given the large sample size (500 women) and the time allocated to the questionnaire (45 to 65 minutes) and the physical examination (15 to 25 minutes), the use of female interviewers at the local level was recommended by the WHO to ensure data reliability. The choice of these interviewers was based on several criteria, mainly on their experience in pre- and post-natal care and data collection

**Table I:**
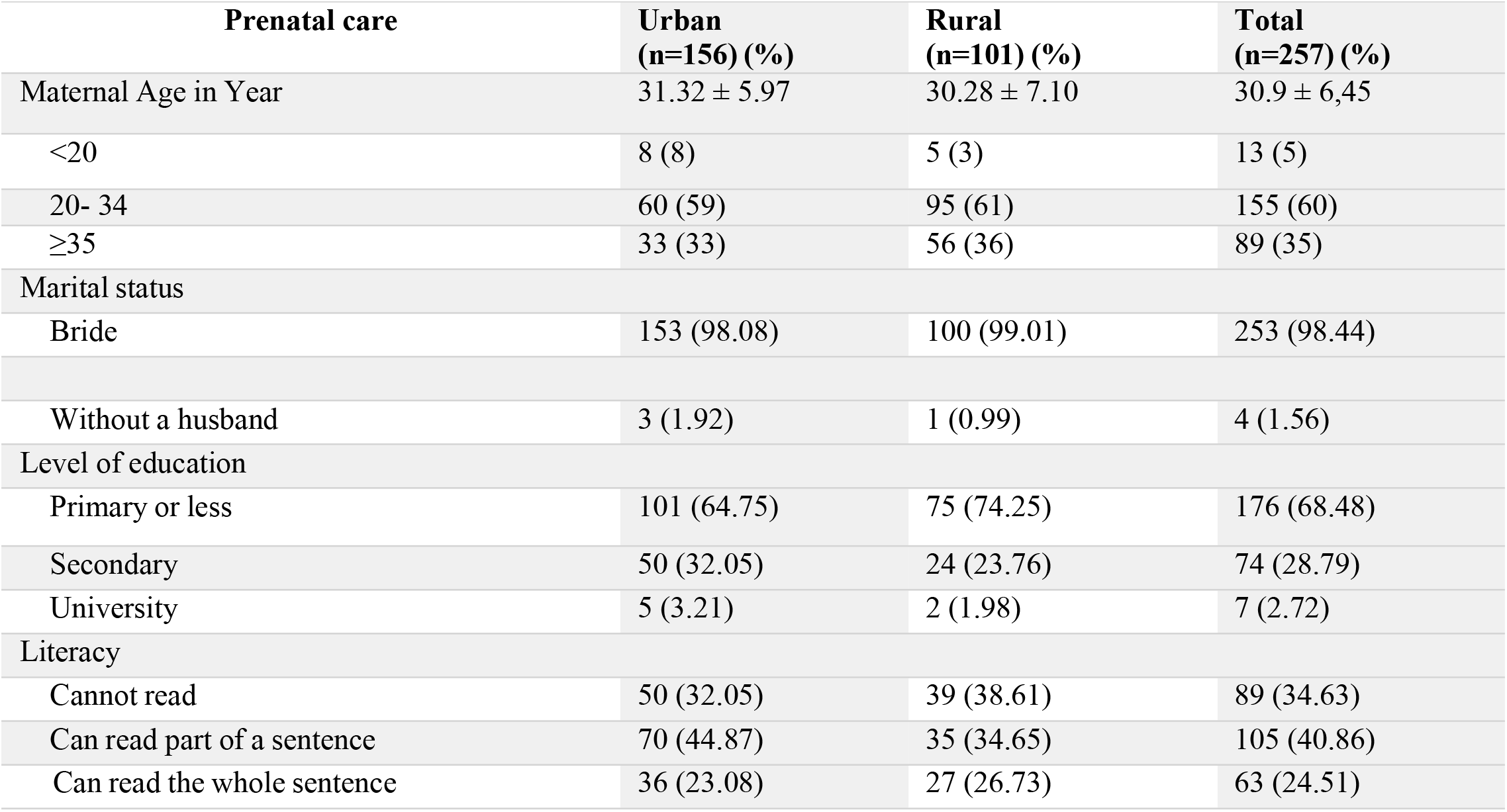
Characteristics of prenatal women:

These interviewers were trained according to WHO guidelines for reliable completion of these tools.

For the administration of the two questionnaires (one for PNC and one for PoNC), each interview with the woman lasted between 15 and 30 minutes in total. The physical examination took between 10 and 15 minutes, so that each woman required a minimum of 25 minutes. In addition to the two questionnaires, a maternal morbidity matrix proposed by the WHO [1] and used in our study was used to operationalize and facilitate the measurement of maternal morbidity. The data analysis was done with the two software programs Epi Info 7 for the quantitative data and Invivo 10 was used for the qualitative data. In addition, for the results of the focus groups, a content analysis was done.

## Results

For both groups of PNC and PoNC women, the average age was 30 years, the proportion of women having their first child was 14.10% -14.43% in both urban and rural groups, compared to those having between 2 and 4 children (76% in both groups). No woman reported a stillbirth. In rural areas (99.01% of ANCs, 97.94% of ANCs) and in urban areas (98.08% of PNC, 100% of PoNC), most women reported being married, compared to only 1.56% of PNC and 2% of PoNC who reported not having a husband. Urban women were the least likely to be illiterate (32.05% in PNC and PoNC vs. 38.61 in PNC and 51.55 in PoNC, rural). While most women were not employed (90.66% in ANC and 94.47% in ANC), the percentage of employed women in urban areas was higher than in rural areas (12.82% in PNC and 8.97% in PoNC, compared to 3.96% in PNC and 3.96% also in PoNC).

**Table II:**
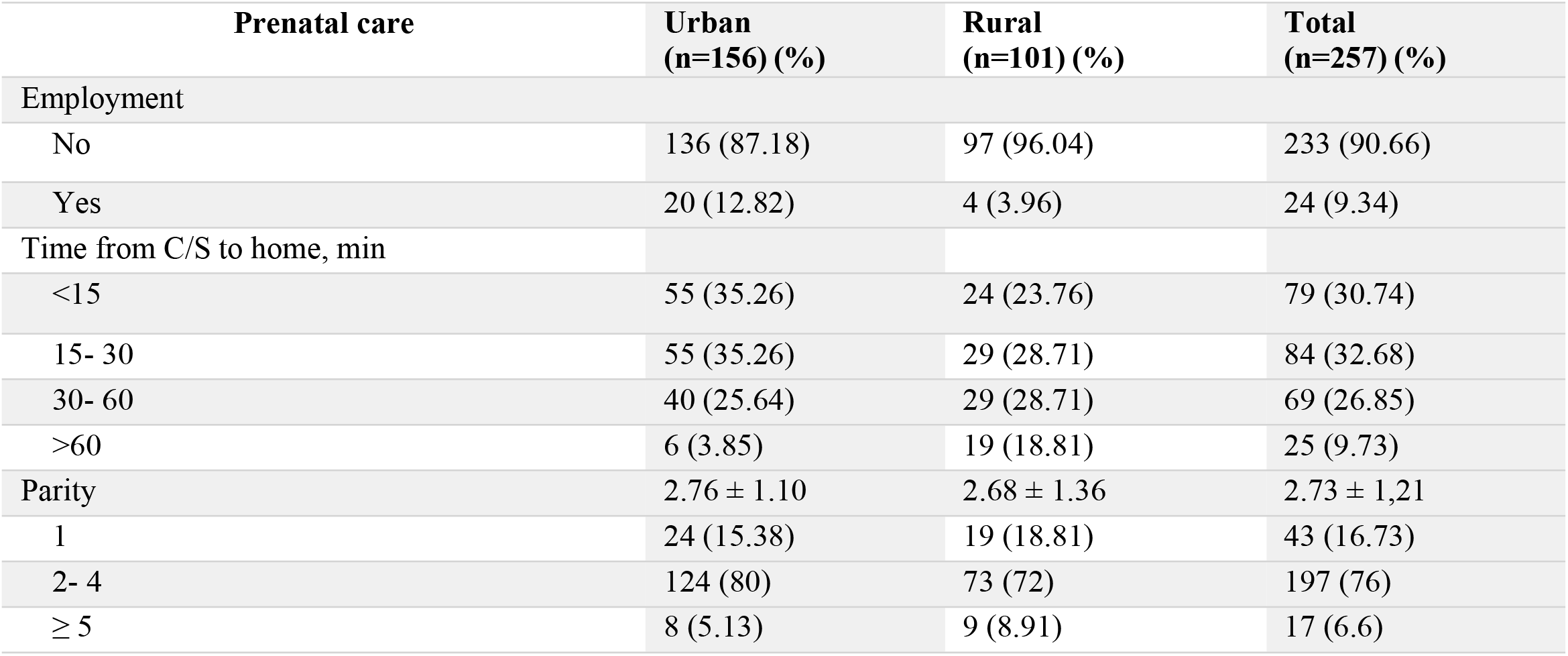
Characteristics of prenatal women (continued):

**Table III:** Characteristics of postnatal women.

| Postnatal care | Urban<br>(n=156) (%) | Rural<br>(n=97) (%) | Total<br>(n=253) (%) |
| --- | --- | --- | --- |
| Employment |  |  |  |
| Maternal Age in Year | 29.78 ± 6.00 | 29.77 ± 6.23 | 29.77 ± 6.08 |
| <20 | 3 (2) | 2 (2) | 5 (2) |
| 20- 34 | 101 (65) | 61 (63) | 162 (64) |
| ≥35 | 52 (33) | 34 (35) | 86 (34) |
| Marital status |  |  |  |
| Bride | 156 (100) | 95 (97.94) | 251 (99.21) |
| Without a husband | 0 (0.00) | 2 (2.06) | 2 (2) |
| Level of education |  |  |  |
| Primary or less | 104 (66.67) | 81 (74.25) | 185 (73.13) |
| Secondary | 43 (27.56) | 14 (14.43) | 57 (22.53) |
| University | 9 (5.77) | 2 (2.06) | 11 (4.35) |
| Literacy |  |  |  |
| Cannot read | 50 (32.05) | 50 (51.55) | 100 (39.53) |
| Can read part of a sentence | 72 (46.15) | 29 (29.90) | 101 (39.92) |
| Can read the whole sentence | 34 (21.79) | 18 (18.56) | 52 (20.55) |
| Employment |  |  |  |
| No | 142 (91.03) | 97 (100) | 239 (94.47) |
| Yes | 14 (8.97) | 4 (3.96) | 14 (5.53) |
| Time from C/S to home, min |  |  |  |
| <15 | 61 (39.10) | 30 (30.93) | 91 (35.97) |
| 15- 30 | 52 (33.33) | 29 (29.90) | 81 (32.02) |
| 30- 60 | 36 (23.08) | 23 (23.71) | 59 (23.32) |
| >60 | 6 (4.49) | 15 (15.46) | 22 (8.70) |
| Parity | 2.76 ± 1.10 | 2.68 ± 1.36 | 2.73 ± 1.21 |
| 1 | 22 (14.10) | 14 (14.43) | 36 (14.23) |
| 2- 4 | 127 (81.41) | 73 (75.25) | 200 (76) |
| ≥ 5 | 7 (4.48) | 10 (10.30) | 17 (6.71) |

### STATUS OF LESS SEVERE MATERNAL MORBIDITIES

#### Self-reported health status of women

Most of the PNC (61.87%) and PoNC (90.91%) women declared that they were in good health, i.e. they had not received any diagnosis from health professionals. Nevertheless, 39.29% in PNC and 37.94% in PoNC declared that their health status was neither good nor bad.

**Table IV:** Health status reported by women during prenatal and postnatal visits.

|  | **Urban**<br>(n=156) | **Rural**<br>(n=101) | **Total**<br>(n=257) |
| --- | --- | --- | --- |
| **Prenatal Consultation** |  |  |  |
| General health status |  |  |  |
| Very good | 0 (0.0) | 5 (4.95) | 5 (1.95) |
| Well | 59 (37.82) | 38 (37.62) | 97 (37.74) |
| Neither good nor bad | 69 (44.23) | 46 (45.54) | 115 (44.75) |
| Bad | 25 (16.03) | 11 (10.89) | 36 (14.01) |
| Very bad | 3 (0.92) | 1 (0.99) | 4 (1.56) |
| Has anyone told you that you have a problem/complication? |  |  |  |
| No | 87 (55.77) | 72 (71.29) | 159 (61.87) |
| Yes | 69 (44.23) | 29 (28.71) | 98 (38.13) |
| Do you have another problem or complaint you want to report? |  |  |  |
| No | 141 (90.38) | 93 (92.08) | 234 (91.05) |
| Yes | 15 (9.62) | 8 (7.92) | 23 (8.95) |
| **Postnatal Consultation** |  |  |  |
| General health status |  |  |  |
| Very good | 3 (1.92) | 2 (2.06) | 5 (1.98) |
| Well | 64 (41.03) | 52 (53.61) | 116 (45.85) |
| Neither good nor bad | 67 (42.95) | 29 (29.90) | 96 (37.94) |
| Bad | 19 (12.18) | 13 (13.40) | 32 (12.65) |
| Very bad | 3 (1.92) | 1 (1.03) | 4 (1.58) |
| Has anyone told you that you have a problem/complication? |  |  |  |
| No | 141 (90.38) | 89 (91.75) | 230 (90.91) |
| Yes | 15 (9.62) | 8 (8.25) | 23 (9.09) |
| Do you have another problem or complaint you want to report? |  |  |  |
| No | 152 (97.44) | 92 (94.85) | 244 (96.44) |
| Yes | 4 (2.56) | 5 (5.15) | 9 (3.56) |

Also, of the 510 women surveyed, only 32 claimed a problem or complaint, or 6.27%. The present study also revealed that the majority of PNC women use medication (80.93%, n=208). These were mainly medicines for the prevention or treatment of anemia or medicines for the treatment of vaginal or urinary infections considered as morbidities.

#### Exposure to risk factors

As part of the definition of morbidity, some contributing factors were explored. We examined exposure to violence by asking women if they had “been afraid of your current or most recent husband or partner” or if “since pregnancy or childbirth, you were pushed, slapped, kicked, or beaten by your husband/partner(s) or someone else? “If women answered yes to any of these questions, they were asked three additional questions related to violence.

Twelve percent (12.23%; n=34) of PNC patients and 20.95% (n=53) of PoNC patients reported being afraid of or experiencing some form of physical violence from their husband/partner(s) or someone else, with rates varying by setting from 12% in urban areas to 29% in rural areas. We also explored substance use reported by women (0.39% - 1.19% overall, Table III). In fact, only four out of 510 women use a substance (tobacco products, alcoholic beverages, cannabis, etc.).

#### Mental health of the women surveyed

In this study, the measurement of mental health of women in PNC and PoNC was based on two tools for measuring anxiety and depression (GAD-7 and PHQ-9). Using both tools, it was revealed that PNC women were more likely to suffer from anxiety (83.65% vs. 29.24% in PNC) and also signs of depression (43% vs. 17.78% in PNC).

**Table V:** Mental health.

|  | **PNC**<br>(n=257) | **PoNC**<br>(n=253) | **Total**<br>(n=510) |
| --- | --- | --- | --- |
| **Anxiety** |  |  |  |
| Yes | 215 (83.65) | 74 (29.24) | 289 (56.66) |
| No | 42 (16.34) | 179 (70.75) | 221 (43.33) |
| **Anxiety Score** | 5.90 ± 4.57 | 1.62 ± 3.36 |  |
| 1- You feel nervous or anxious? | 215 (83.65) | 74 (29.24) | 289 (56.66) |
| 2- Not being able to stop or control your anxiety | 32 (12.45) | 12 (4.74) | 44 (8.62) |
| 3- Worrying about many different things different | 86 (33.46) | 24 (9.48) | 110 (21.56) |
| 4- Relaxation disorder | 34 (13.22) | 12 (4.74) | 46 (9.01) |
| 5- Being so restless that it is difficult to sit still | 52 (20.23) | 19 (7.50) | 71 (13.92) |
| 6- Become easily irritable | 215 (83.65) | 65 (25.69) | 280 (54.90) |
| 7- To be afraid as if something terrible could happen | 125 (48.63) | 21 (8.30) | 146 (28.62) |
| No | 154 (98.72) | 96 (98.97) | 250 (98.81) |
| Yes | 2 (1.28) | 1 (1.03) | 3 (1.19) |
| **Exposure to violence** |  |  |  |
| No | 132 (84.62) | 68 (70.10) | 200 (79.05) |
| Yes | 24 (15.38) | 29 (29.90) | 53 (20.95) |

These two mental health measurement tools showed that 14% (n=36) in PNC and 17.78% (n=45) in PoNC have a score that exceeds 10. This means that they present one or more signs of psychiatric disorders. It also appears from the results of this study that the signs of anxiety that were most raised by the women were the first (56.66%, n=289) and the sixth (irritability) (54%, n=280).

##### Obesity and Sexual Satisfaction

Given the global prevalence of obesity, women’s height and weight were also documented (Table VII). Of the 510 women interviewed, 178 had a body mass index greater than or equal to 30. There were no differences between urban and rural areas or between the two groups of women (PNC and PoNC).

**Table VI:**
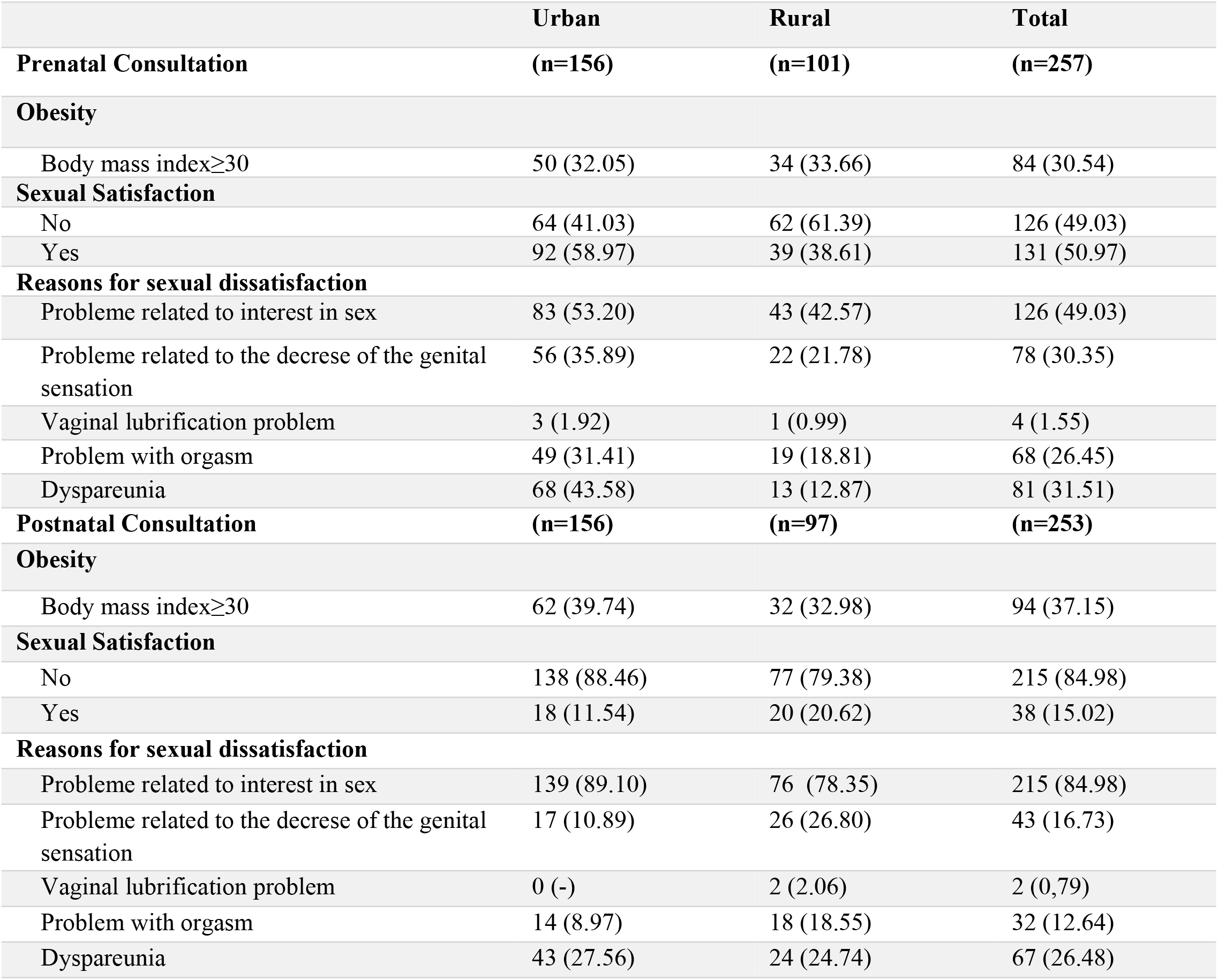
The frequency of Obesity and sexual satisfaction during the prenatal and postnatal consultation.

|  | Urban | Rural | Total |
| --- | --- | --- | --- |
| **Prenatal Consultation** | **(n=156)** | **(n=101)** | **(n=257)** |
| **Obesity** |  |  |  |
| Body mass index$\geq 30$ | 50 (32.05) | 34 (33.66) | 84 (30.54) |
| **Sexual Satisfaction** |  |  |  |
| No | 64 (41.03) | 62 (61.39) | 126 (49.03) |
| Yes | 92 (58.97) | 39 (38.61) | 131 (50.97) |
| **Reasons for sexual dissatisfaction** |  |  |  |
| Probleme related to interest in sex | 83 (53.20) | 43 (42.57) | 126 (49.03) |
| Probleme related to the decrease of the genital sensation | 56 (35.89) | 22 (21.78) | 78 (30.35) |
| Vaginal lubrication problem | 3 (1.92) | 1 (0.99) | 4 (1.55) |
| Problem with orgasm | 49 (31.41) | 19 (18.81) | 68 (26.45) |
| Dyspareunia | 68 (43.58) | 13 (12.87) | 81 (31.51) |
| **Postnatal Consultation** | **(n=156)** | **(n=97)** | **(n=253)** |
| **Obesity** |  |  |  |
| Body mass index$\geq 30$ | 62 (39.74) | 32 (32.98) | 94 (37.15) |
| **Sexual Satisfaction** |  |  |  |
| No | 138 (88.46) | 77 (79.38) | 215 (84.98) |
| Yes | 18 (11.54) | 20 (20.62) | 38 (15.02) |
| **Reasons for sexual dissatisfaction** |  |  |  |
| Probleme related to interest in sex | 139 (89.10) | 76 (78.35) | 215 (84.98) |
| Probleme related to the decrease of the genital sensation | 17 (10.89) | 26 (26.80) | 43 (16.73) |
| Vaginal lubrication problem | 0 (-) | 2 (2.06) | 2 (0.79) |
| Problem with orgasm | 14 (8.97) | 18 (18.55) | 32 (12.64) |
| Dyspareunia | 43 (27.56) | 24 (24.74) | 67 (26.48) |

In addition, the results of the present survey showed that the majority of the women interviewed suffered from sexual dissatisfaction, 49.03% (n=126) in PNC and 84.98% (n=215) in PoNC. This problem was more pronounced among PoNC women and especially among PoNC women in urban areas (88.46%, n=138).

#### The main complications diagnosed during PNC and PoNC

The maternal morbidity matrix proposed by the WHO and used in this study made it possible to operationalize and facilitate the measurement of maternal morbidity. Indeed, the morbidities diagnosed by the health professionals (FH and GP) were summarized according to the guidelines of this matrix.

**Table VII:**
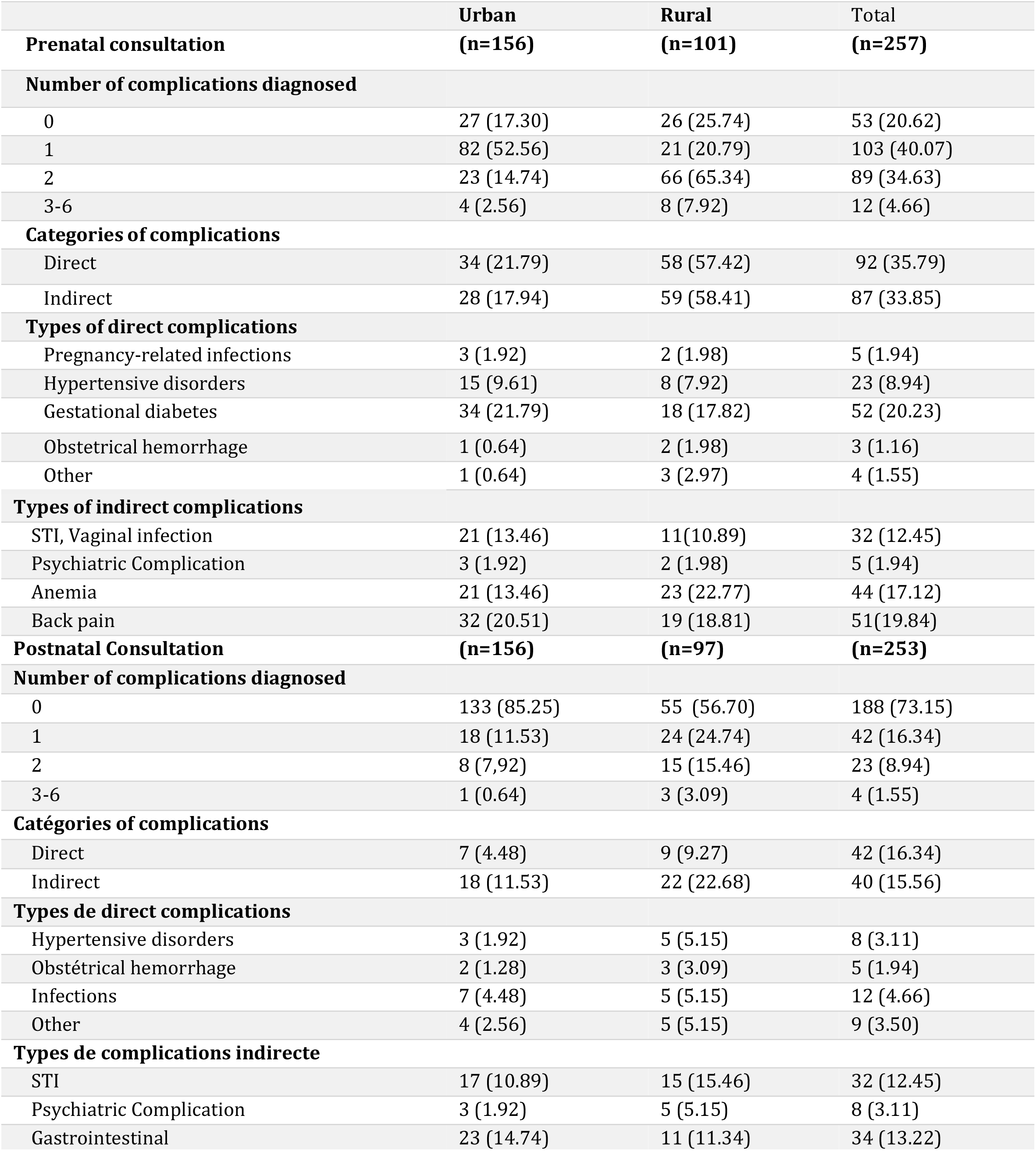
The main complications diagnosed.

Referring to the WHO pilot study, direct obstetric complications were grouped into hypertensive disorders, obstetric hemorrhage, pregnancy-related infections, and other specific direct conditions, while indirect complications were classified into cardiovascular, hematologic, endocrine, and other complications.

The major direct and indirect complications diagnosed in this study are presented in the table above. From this table, it appears that the majority of pregnant women, who participated in the test (79.38%), were diagnosed with one or more morbidities (including depression or anxiety screening). Of the 257 pregnant women, only 20.62% were healthy.

The high number of diagnosed complications was significantly reduced in the postpartum period. In fact, most postpartum women (73.15%) were healthy and only 25.28% had one or two diagnosed complications. We also diagnosed PNC women with direct (35.79%, n= 92) or indirect (33.85%, n= 87) complications, whereas PoNC women were more likely to be diagnosed with one or more indirect morbidities (15.56%, n=40), than with direct complications (6.32%, n=16).

Using the direct complication groups of the WHO maternal morbidity matrix, women generally with PNC had a high rate of gestational diabetes (20.23%, n=52), followed by hypertensive disorders (8.94%, n=23). The category of hypertensive disorders combines preeclampsia, gestational hypertension, and clinical findings of elevated blood pressure. Clinical examination revealed five women with depression in ANC and eight cases in PNC. No women were diagnosed with anxiety by a general practitioner).

## Discussion

The present study consists in testing the measurement tools of maternal morbidity proposed by the WHO [1] and in describing the situation of less serious maternal morbidities at the level of the Primary Health Care facilities in the prefecture of Marrakech. It revealed a discrepancy between, on the one hand, the complaints expressed by women at the prenatal and postnatal consultation, and on the other hand, the clinical morbidity diagnosed by the physician. Indeed, almost half of the women (47% in PNC and 49% in PoNC) declared that they were in good health. However, the majority of pregnant women, 79.36%, were diagnosed with at least one morbidity compared to only 26.83% in postnatal. For prenatal women, the first category of diagnosis was related to gestational diabetes (20.23%), the second was represented by anemia (17.12%), while hypertensive disorders and mental health related disorders were diagnosed in only 8.94% and 1.94% respectively. For gestational diabetes, according to a case-control study carried out at the level of the PHSS in the prefecture of Marrakech between 2016 and 2017, the prevalence of gestational diabetes is 23.7% (Utz et al, 2017). This result corroborates that of our study. In addition, hypertensive disorders were among the most frequent diagnoses raised by the WHO pilot study [10]. For hypertensive disorders in pregnancy, a Ghanaian study [15] documented a prevalence of 11.3%, a much higher rate than this study.

The present study also showed that the prevalence of violence varies considerably among women. However, the rates were generally higher (12% to 14%) than those in the Malawi study (8.4%), and lower than the results of the Kenya study (17.4%). In contrast, the rate of violence in Jamaica was similar (12%). It was found by the interviewers that women responded less to the question about violence in general. However, when asked specific questions about their experience of domestic violence, they were more likely to respond very specifically. It is therefore relevant to address exposure to violence, particularly in domestic violence, as it is related to poor utilization of prenatal care [16], low birth weight and preterm delivery [17], postpartum depression [18], and even pregnancy-related suicide or homicide [19]. In addition, our study found that the prevalence of emotional abuse was more frequent than physical abuse. Studies corroborate these results by also pointing out that women consider this situation normal, for them moral violence and conflicts are part of conjugal life [17, 20]. The results of our study also show that there is a difference in the prevalence of domestic violence between urban and rural areas. This result does not corroborate with a study conducted in Egypt in 2017 [18], which showed that women in rural areas are more exposed to domestic violence than those in urban areas. In our study, self-reported depression and anxiety (using GAD-7 and PHQ-9 for screening) showed a high prevalence of mental disorders (14% in PNC and 17.78% in PoNC) compared to the WHO pilot study [10] which revealed rates of 6.3% for PNC and 2.2% for PoNC. According to several studies including this one, many social and economic factors have been associated with prenatal and postnatal depression, including first pregnancy [21], and the case of domestic violence [22]. According to a study published in 2016 [23], identifying and treating these women is critical not only for their health, but also for the survival and development of their children. It is then recommended that prenatal and postnatal care be improved to ensure that these women receive appropriate care [10]. The results of our study corroborate those of two other studies [24, 25] on sexuality during pregnancy and the postpartum period. The first one involved 570 pregnant women, interviewed at T1 (Fifth month of pregnancy), T2 (at 1 month postpartum), T3 (at 4 months postpartum) and T4 (at 12 months postpartum), and showed that at T1 and T2, the majority of women showed significantly less sexual activity and less sexual satisfaction. The second longitudinal cohort study just published in 2018 on 832 prenatal and postpartum women confirms this evidence. Indeed, almost half of the women (46.3%) reported a lack of interest in sexual activity, 43% experienced a lack of vaginal lubrication and 37.5% of the women included had dyspareunia 6 months after birth. The authors of both studies, suggest that practitioners provide family-centered maternity care, they should counsel couples on typical patterns of sexuality during pregnancy and postpartum, and on usual patterns during breastfeeding. Accurate information can help couples feel more comfortable during the transition periods before and after delivery. A discussion of expected changes in sexuality should be routinely introduced during prenatal care.

## Strengths and limitations of the study

### Study strengths

Our study is the first in North Africa to test the measurement tools for less severe morbidity proposed by the WHO [1]. It is the third in Morocco to address the topic of maternal morbidity and the first to address the measurement of less severe maternal morbidity in Morocco. Both women and interviewers and other health professionals and partners measure the relevance of the study through the acceptance of participation in the study.

### Study Limitations

Time and resource constraints were a major limitation to our study, preventing us from validating the measurement tools tested with regional and central level officials. The time constraint also limited us from making associations between certain variables and from integrating other populations such as husbands and family, NGOs, and other partners. Another limitation is the coordination with general practitioners in order to diagnose morbidities and to follow up on women in order to describe the evolution of morbidities.

## Recommendations

Based on the results of the present study as well as on the WHO recommendations for interventions to combat maternal morbidity, avenues for improvement are proposed [26].

### Paradigm shift

#### Pregnancy should be seen as an opportunity

Pregnancy should be viewed as an opportunity to improve women’s overall health, so that quality maternal health care can ensure benefits for the current pregnancy, future pregnancies, and long-term health. The conceptualization of pregnancy thus situates maternal health across the life course and provides women with health services in a seamless and integrated manner, ranging from family planning to pre-pregnancy care, pregnancy, labor, delivery, and the postpartum period, as well as non-communicable and reproductive health care. Pregnancy is unique as an event in a woman’s life. It may reveal underlying or undiagnosed, dormant or unrecognized conditions. Pregnancy then provides an opportunity to identify these women and intervene to reduce the longer-term consequences. Merging maternal health and non-communicable disease strategies.

Maternal health care has traditionally focused on the diagnosis and management of obstetric complications. Maternal health is closely linked to non-communicable diseases. Pregnancy complications can increase the prevalence of chronic health problems, affecting not only future pregnancies but also women’s long-term health [27, 28]. Non communicable diseases combined are the leading cause of death among women worldwide, accounting for 65% of all deaths [29]. Health services are the only potential points of contact for diagnosis and prevention of NCDs. Pregnant and postpartum women and NCD programs can no longer be addressed separately. This is reflected in the new strategy for the control of preventable deaths.

#### Expanding the reach of maternal health services

The need to better understand the role of maternal morbidity in women’s health implies a greater appreciation of what these complications do to women’s health and well-being and may contribute to the negative experiences of pregnancy itself. A key element of the present study is that indirect or medical complications contributed significantly to maternal morbidity. To address this, the maternal and newborn health strategy must extend beyond pregnancy and childbirth to include links to premarital care and long-term health. Premarital counseling should be considered the “third” routine component of maternal health care, and of equal importance to prenatal and postnatal care. Similarly, emphasis should be placed on strengthening postpartum services, extending postpartum care beyond six weeks if necessary, and improving integration with other medical specialties. Postpartum care could be seen as an opportunity to promote women’s health. With all of the actions currently being implemented by the Ministry of Health, maternal mortality will continue to decline, meaning that the number of women who escape will increase and women will have more morbidity as a result.

#### Adopting a rights-based approach

By adopting a rights-based approach to health and well-being, women can reach their full potential, and women and newborns will not only survive but also thrive. This requires a comprehensive approach that also includes more distal, non-clinical risk factors or social determinants of health. These factors not only create social vulnerabilities, but also influence health behavior and access to care, which requires a rights-based approach to maternal health. In this sense, the following is recommended :

- Implement strategies to address women’s social vulnerability, which can be particularly acute for young, unemployed or unmarried women. These women are at high risk for domestic violence, anxiety and depression, malnutrition, and limited access to health care.
- Enhance intersectorality and take a community-based approach to address the social determinants of health. Involve the husband and immediate family because of the important role they play in providing emotional and financial support.
- Ensure free or fully covered essential maternal and newborn health services.

### Translating paradigms into interventions

#### Premarital consultation

The premarital period is a critical entry point for hygiene, nutrition, and childbirth preparation. Adolescent girls, for whom targeted interventions are needed during this period, are particularly vulnerable and neglected [30].

First, engaging women of childbearing age during the pre-pregnancy period helps determine fertility intentions and thus allows for more careful and thoughtful pregnancy planning, especially when women seek contraceptive services. Second, evidence increasingly points to pre-pregnancy care to improve women’s health and maternal and newborn outcomes [31, 32]. Counseling is particularly important for women with preexisting medical complications. Some medical complications may be accentuated by the physiological changes of pregnancy, and close monitoring of a carefully planned pregnancy is possible.

#### Prenatal consultation

According to our study as well as other studies on maternal morbidity, some pre-existing complications are diagnosed for the first time during pregnancy (such as diabetes and hypertension), a general health assessment could be included in the maternal history taking. The findings of the pilot study also point to the possibility of expanding the scope of PNC beyond chronic disease management to include a simple method of screening for domestic violence and mental health. WHO has long recognized that integrating antenatal care with other health services is a key strategy for reducing missed opportunities for patient contact and for effectively addressing the overall health needs of women.

ANC has traditionally been associated with prevention of mother-to-child transmission services and syphilis screening. Countries can capitalize on ANC coverage by further integrating chronic disease services.

#### The post-natal consultation

The postpartum period remains a time that is not given the same importance by policymakers and health professionals as prenatal care and delivery. Once a woman and her newborn are rescued during delivery, there is no pressure on providers to know the woman’s fate [33]. However, the results of the present study identified that postpartum follow-up is particularly important to address symptoms and complications that women identify as troublesome and impacting their functioning. Postpartum women expressed that the range of experiences they encounter is not fully recognized and that the care offered is limited to obstetrical observations. It is recommended that:

Provide more comprehensive care by addressing issues of sexuality, mental health, domestic violence, physical recovery from childbirth, contraception, emotional well-being….

Capitalize on women’s presence in health facilities after delivery to meet their needs and those of their newborns. Provide psychosocial support by a trained individual for the prevention of postpartum depression in women at high risk of developing this complication. Psychosocial and psychological interventions have been shown to reduce the number of women at risk who develop postpartum depression [34]. These interventions include the provision of intensive, professional postpartum home visits, telephone peer support and interpersonal psychotherapy.

## Conclusion

As the international community focuses on reducing maternal mortality, there is an urgent need to define and measure maternal morbidity. Beyond 2015, this is an investment that no one can ignore. All countries must now move beyond survival to establish integrated health services that maximize women’s health, well-being, and potential across the life course [26]. This reflects the full value of women as members of families, communities, societies and economies. This study supports the added value of maternal morbidity measurement tools to meet information needs, and comprehensive and integrated management that is part of the continuum of care. Although women interviewed report a high level of satisfaction with MM tools, and despite growing support for these tools from health professionals, managers, and international partners, their adoption may take time. Reasons include limited available resources, lack of time to complete the tools as proposed by WHO and buy-in from all health professionals. Based on the results of this study, we have proposed to the key actors ways to improve, which consists first of all in adopting a broader approach to maternal health that responds to contemporary challenges and the most frequent complications. This means moving beyond traditional models of maternity care where women are in contact with health systems only during pregnancy until 6 weeks after delivery. The provision of maternal health services can have a broader scope throughout a woman’s life cycle, including premarital care, longer-term health care, and better integration of existing health programs and services. Maternal health and communicable and non-communicable disease programs should be synergistic. The proposed recommendations, as well as the simplified form to be integrated into the pregnancy and postpartum surveillance form already in place at the PHSS level, will help provide evidence-based and comprehensive data on maternal morbidity and therefore facilitate decision-making. Investing in maternal morbidity will not only prevent maternal and neonatal deaths, but also contribute to the well-being of the mother and her newborn and thus achieve MDG 3 by 2030.

## Data Availability

All relevant data are within the manuscript

## ABBREVIATIONS

PoNC: (Postnatal consultation)
PNC: (Prenatal consultation)
GD: (General Doctor)
PHCN: (Primary health care Network)
PWCP: (Pregnant woman care program)
OMM: (Measuring maternal morbidity)
WHO: (Word Health Organization)

## DECLARATIONS

Ethics approval and consent to participate

The research protocol was approved by the Ethics Committee for Bimedical Research under the Faculty of Medicine and Pharmacy of Rabat. All study participants signed a consent form before the start of the study.

### Consent for publication

You will find in attachments the consents for the publication of the two authors.

### Availability of data and material

All the database and material used in this study are available.

We confirm that all methods were performed in accordance with the relevant guidelines and regulations.

If someone wants to request the data from this study, he can contact

### Authors’ contributions

Hanane Hababa searched the literature, extracted data, synthesized data and developed the first draft of the manuscript; Bouchra Assarag carefully checked the manuscript; To provided essential methodological advice. The authors have read and approved the final manuscript.

## Acknowledgements

We would like to express our deep gratitude to all the participants in this study, parturient, midwives and nurses, managers and officials at the level of the Ministry of Health.

---

**Figure 1:**
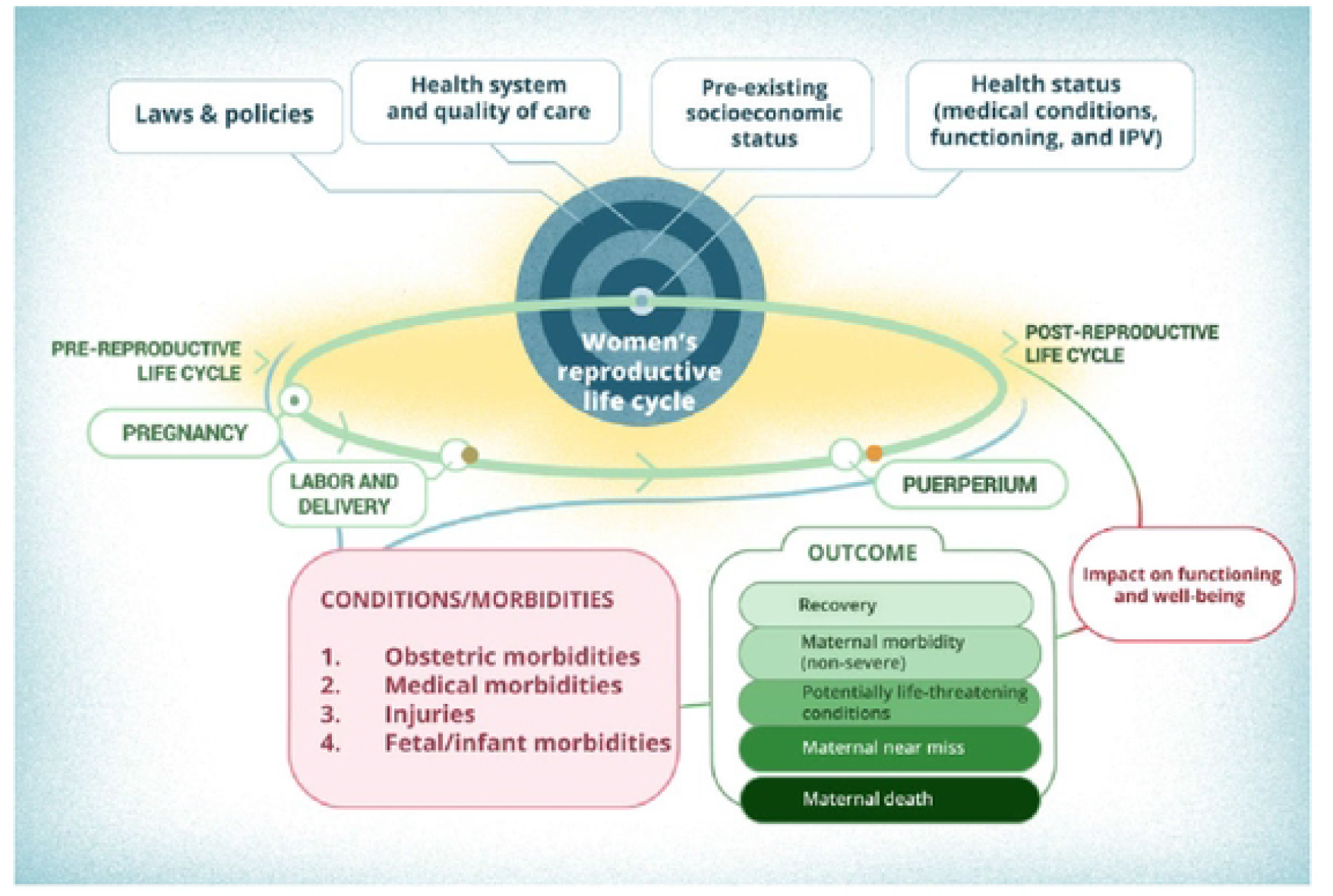
Conceptual framework for measuring maternal morbidity (MMM) [12].

## Références

[1] Say L, Barreix M, Chou D, et al. Maternal morbidity measurement tool pilot: Study protocol. Reprod Health. 2016;13:69.

[2] Graham W, Woodd S, Byass P, et al. Diversity and divergence: the dynamic burden of poor maternal health. (2016). Lancet;388:2164–75

[3] Singh S, Darroch JE, Ashford LS. Adding It Up: The Costs and Benefits of Investing in Sexual and Reproduc ve Health 2014. New York: Gu macher Ins tute; 2017.

[4] Knaul F, Langer A, Atun R, Rodin D, Frenk J, Bonita R. Rethinking maternal health. Lancet Glob Health. 2016;4:e227– e228.

[5] Chou D, Tunçalp Ö, Firoz T, et al. Constructing maternal morbidity - towards a standard tool to measure and monitor maternal health beyond mortality. (2016). BMC Pregnancy Childbirth;16:45

[6] Kazuyo; M et al. Consequences of maternal morbidity on health-related functioning: a systematic scoping review. (2017).

[7] Fortney J, Smith J. Measuring maternal morbidity. In: Berer M, Ravindran Tks, eds. Safe Motherhood Initiatives: Critical Issues. London: Reproductive Health Matters; 1999:43–50.

[8] Hardee K, Gay J, Blanc A. Maternal morbidity: Neglected dimension of safe motherhood in the developing world. Glob Public Health. 2012 ;7:603–617.

[9] Haddad SM, Ceca JG, Parpinelli MA, et al. From planning to practice: Building the national network for the surveillance of severe maternal morbidity. BMC Public Health. 2011;11:283.

[10] Barreix M, Barbour K, McCaw-Binns A, et al. Standardizing the measurement of maternal morbidity: Pilot study results. Int J Gynecol Obstet. 2018;141(Suppl.1):10–19.

[11] Vanderkruik RC, Tuncalp O, Chou D, Say L. Framing maternal morbidity: WHO scoping exercise. BMC Pregnancy Childbirth. 2013; 13:213.

[12] Filippi V, Chou D, Barreix M, Say L. A new conceptual framework for maternal morbidity. Int J Gynecol Obstet. 2018;141(Suppl.1):4–9.

[13] Barbour, KD et al. Developing maternel morbidity identification algorithms : results from the pilot study of the WHO Maternel Morbidity Measurement Tool. (2017).

[14] World Health Organization. WHO Disability Assessment Schedule 2.0 (WHODAS 2.0). Geneva: WHO; 2017.

[15] Browne JL, Vissers KM, Antwi E, et al. Perinatal outcomes after hypertensive disorders in pregnancy in a low resource se ng. Trop Med Int Health. 2015;20:1778–1786.

[16] Singh JK, Evans-Lacko S, Acharya D, Kadel R, Gautam S. Intimate partner violence during pregnancy and use of antenatal care among rural women in southern Terai of Nepal. Women Birth. 2017;pii: S1871–5192(16)30143–3. [Epub ahead of print].

[17] Bailey BA. Partner violence during pregnancy: Prevalence, effects, screening, and management. Int J Womens Health. 2010;2: 183–197.

[18] El-Hosary EA, Amany A, Emaghawry Eldeeb M. Effect of Domestic Violence on Pregnancy Outcomes among Rural and Urban Women. 2017 ; IOSR Journal of Nursing and Health Science Volume 6, Issue 3 Ver. II (May. - June. 2017), PP 35–42

[19] Palladino CL, Singh V, Campbell J, Flynn H, Gold K. Homicide and suicide during the perinatal period: Findings from the National Violent Death Reporting System. Obstet Gynecol. 2011;118:1056–1063.

[20] Villas Boas Teixeira S, Aparecida Vasconcelos Moura M, Rangel da Silva L, Beatrix Azevedo Queiroz A, Ventura de Souza K, Albuquerque Netto L. Intimate partner violence against pregnant women: the environment according to Levine’s nursing theory. 2015 Journal of school of Nursing, USP.

[21] Tachibana Y, Koizumi T, Takehara K, et al. Antenatal risk factors of postpartum depression at 20 weeks gesta on in a Japanese sample: Psychosocial perspectives from a cohort study in Tokyo. PLoS ONE. 2015;10:e 0142410.

[22] Gausia K, Fisher C, Ali M, Oosthuizen J. Antenatal depression and suicidal idea on among rural Bangladeshi women: A community-based study. Arch Womens Ment Health. 2009;12:351–358.

[23] Surkan PJ, Patel SA, Rahman A. Preventing infant and child morbidity and mortality due to maternal depression. Best Pract Res Clin Obstet Gynaecol. 2016; 36:156–168.

[24] Hyde et al, (1996). Sexuality during Pregnancy and the Year postpartum

[25] O’Malley D, Agnes Higgins A, Begley C, Daly D, Smith V. Prevalence of and risk factors associated with sexual health issues in primiparous women at 6 and 12 months postpartum; a longitudinal prospective cohort study (the MAMMI study). 2018. MC Pregnancy and Childbirth (2018) 18:196

[26] Firoz T, McCaw-Binns A, Filippi V, et al. A framework for healthcare interventions to address maternal morbidity. Int J Gynecol Obstet. 2018;141(Suppl.1):61–68.

[27] Feig DS, Zinman B, Wang X, Hux JE. Risk of development of diabetes mellitus after diagnosis of gestational diabetes. CMAJ. 2008;179:229–234.

[28] Herath H, Herath R, Wickremasinghe R. Gestational diabetes mellitus and risk of type 2 diabetes 10 years after the index pregnancy in Sri Lankan women – a community based retrospective cohort study. PLoS ONE. 2017;12:e0179647.

[29] World Health Organization. The Global Burden of Disease: 2004 Update. Geneva: WHO; 2008.

[30] Bhutta ZA, Lassi ZS, Bergeron G, et al. Delivering an action agenda for nutrition interventions addressing adolescent girls and young women: Priorities for implementation and research. Ann N Y Acad Sci. 2017;1393:61–71.

[31] Dean SV, Imam AM, Lassi ZS, Bhu a ZA. Systematic Review of Preconception Risks and Interventions.http://mother-childlink.tghn.org/site_media/media/articles/Preconception_Report.pdf. Accessed December 18, 2017.

[32] Johnson K, Posner SF, Biermann J, et al. Recommendations to improve preconception health and health care – United States. A report of the CDC/ATSDR Preconception Care Work Group and the Select Panel on Preconception Care. MMWR Recomm Rep. 2006; 6:1–23.

[33] Assarag, B (2015). La morbidité maternelle sévère aigue au Maroc - Facteurs explicatifs et conséquences : une base d’évidence nécessaire pour une réponse appropriée

[34] Dennis CL, Dowswell T. Psychosocial and psychological interventions for preventing postpartum depression. Cochrane Database Syst Rev. 2013;(2):CD001134.

